# Do human recordings reveal drastic modulations in the discharge of striatal projection neurons in Parkinson’s disease?

**DOI:** 10.1101/2020.03.04.20030999

**Authors:** Dan Valsky, Zvi Israel, Thomas Boraud, Hagai Bergman, Marc Deffains

## Abstract

Dopamine depletion of the striatum plays a key role in the pathophysiology of Parkinson’s disease (PD), but our understanding of the changes in the discharge rate and pattern of the striatal projection neurons (SPNs) remains limited. Here, we recorded multi-unit signals from the striatum of PD (N = 934) and dystonic (N = 718) patients undergoing deep brain stimulation surgeries. Using an innovative automated data-driven approach to classify striatal units, we showed that the SPN discharge rate is inversely proportional to the isolation quality and stationarity of the SPNs. In contrast to earlier studies in both PD patients and the non-human primate model of PD, we found no drastic changes in the spiking activity (discharge rate and pattern) of the well-isolated and stationary SPNs of PD patients compared to either dystonic patients or the normal levels of striatal activity reported in healthy animals. Moreover, cluster analysis using SPN discharge properties did not characterize two well-separated SPN subpopulations. There was therefore no specific SPN subpopulation (D1 or D2 SPNs) strongly affected by the pathological state. Instead, our results suggest that moderate changes in SPN discharge are most likely amplified by basal ganglia downstream structures, thus leading to the clinical (motor and non-motor) symptoms of PD.

**Significance statement:** In Parkinson’s disease (PD), the loss of the midbrain dopaminergic neurons leads to massive striatal dopamine depletion that provokes abnormal activity throughout the basal ganglia. However, the impact of dopamine depletion on neuronal activity in the striatum is still highly debated. We recorded and examined the neuronal activity in striatum of PD and dystonic patients undergoing deep brain stimulation surgeries. We found that striatal activity was not drastically higher in PD patients compared to either dystonic patients or the normal levels of striatal activity reported in animal studies. In PD, moderate changes in striatal basal activity are therefore most likely amplified by basal ganglia downstream structures.

## Introduction

Parkinson’s disease (PD) and dystonia are two of the most common movement disorders, and have a wide spectrum of etiologies and clinical presentations. To date, the pathophysiology of PD and dystonia is still debated. Unlike in PD, there is no degeneration of the midbrain dopaminergic neurons in dystonia. Nevertheless, an imbalance between the midbrain dopaminergic and striatal cholinergic systems (1–4), as well as cerebellum dysfunction, (5–9) are present in both PD and dystonia. Moreover, both diseases are traditionally viewed as basal ganglia (BG) disorders (10, 11) and deep brain stimulation (DBS) in BG (subthalamic nucleus, STN and internal segment of the globus pallidus, GPi) is an effective invasive treatment for both diseases (12–15).

The striatum (i.e., the main input structure of the BG network) is the main recipient of midbrain dopaminergic neurons (16) and also receives major cerebellar projections (17, 18). Most (>95%) of the striatal neurons are medium spiny projection neurons (SPNs) that receive afferents from the cortex and the thalamus, and together with the STN, innervate the central (i.e., external segment of the globus pallidus, GPe) and output (i.e., GPi and substantia nigra reticulata, SNr) BG structures (19, 20). Therefore, alteration of striatal signaling disrupts normal BG activity and may lead to the manifestation of the motor and non-motor symptoms of PD and dystonia (19–21).

Dysregulation of BG activity may consist of changes in discharge rate (19, 20). Unlike in all other BG structures, extracellular recordings of spiking activity in non-human primates (NHPs) reveal that SPNs have a very low discharge rate (∼1-2Hz at rest) and are phasically active (i.e. emit short bursts) around relevant behavioral events (22–24). However, earlier studies of the BG in the NHP model of PD have mainly focused on the STN, GPe and GPi which are structures with a high frequency tonic discharge (i.e. 25-70Hz at rest) (25–27). These studies have reported excessive GPi/SNr inhibitory inputs in PD leading to an increase of BG inhibitory outputs to the thalamus and the frontal cortex motor areas (27). Conversely, it is assumed that dystonia (and other hyper-kinetic states) are characterized by reduced GPi/SNr activity (28).

More recent studies of the BG in animal (rodent and NHP) models of PD and human patients (undergoing DBS procedures) have focused on changes in discharge patterns and synchronization. Parkinsonism-related β oscillations have been observed in local field potentials (LFPs) recorded in all BG structures, including the striatum (25, 29–31). Similarly, low frequency (4-12Hz) LFP oscillations have been recorded in the BG network of dystonic patients (32, 33). Finally, synchronous β oscillations are commonly observed in the spiking activity of the STN, GPe and GPi of MPTP-treated monkeys (25, 34, 35) and PD patients (36, 37).

Nevertheless, direct evidence for abnormal activity of the striatal SPNs is still elusive. Earlier studies in the NHP model of PD reported striking increases (∼15-fold increase from the normal discharge rate of ∼1-2Hz) in the firing rate of the SPNs subsequent to striatal dopamine depletion and the induction of parkinsonism (38, 39). The same research group also reported a high discharge rate of SPNs recorded in PD and dystonic patients (∼30Hz and 9Hz, respectively) (39). They also found a significant change in the firing pattern of striatal neurons, with many SPNs exhibiting bursting activity in PD patients and MPTP monkeys as compared to a smaller fraction in patients with dystonia (39). Finally, the SPNs of patients with an essential tremor (ET, considered as non BG disorder) have a very low discharge rate (∼2Hz) and no tendency to burst, as reported in normal NHPs (38–40).

These spectacular changes in the discharge rate and pattern of SPNs in the NHP model of PD run counter results obtained in our research group. We recorded the activity of SPNs and other BG neurons in Vervet monkeys before and after systemic 1-methyl-4-phenyl-1,2,3,6-tetrahydropyridine (MPTP) treatment and the induction of severe parkinsonian symptoms. Although we found robust changes in discharge properties (rate, pattern and synchronization) of the STN, GPe and BG output structures, we did not observe any difference in the discharge rate (∼2-3Hz) or pattern of SPNs in the MPTP-treated monkeys in comparison to the recordings in the same monkeys before MPTP treatment (25, 34, 41).

Extracellular recordings of SPN spiking activity of anesthetized (42–44) and awake (42, 45) rats before and after striatal dopamine depletion by 6-hydroxydopamine (6-OHDA) treatment have revealed a significant, but very slight, increase in the SPN discharge rate. Rodent studies make it possible to differentiate between SPNs expressing D1 and D2 dopamine receptors. A significant imbalance, [but see (46)] in the discharge rate and calcium dynamics of D1 and D2 SPNs was observed in the dopamine-depleted striatum (43, 44, 47). In particular, D2 SPNs increased their discharge (43, 44) and were also prone to being entrained to parkinsonian β oscillations (44). Nevertheless, the absolute increase in the discharge rate even of the D2 SPNs was still modest (from ∼0.5 to ∼2.8Hz) (44). Moreover, recent studies have reported no significant increase in the low discharge rate of either SPN subpopulation in striatal dopamine-depleted mice (46, 48).

To date, only one research group has been able to record striatal spiking activity in patients (39). Accordingly, we examined our human patient data to check whether we could observe similar drastic spiking activity increases in striatum of PD patients. Unlike in animal (rodent and NHP) studies, there is no control condition (healthy recordings). Furthermore, extracellular recording methods do not allow us to discriminate between the spiking activity of the striatal D1 and D2 SPNs. Nevertheless, the current study was conducted under the assumption that if only one population of striatal neurons is strongly affected by the PD state, we should observe a significantly higher discharge rate of SPNs than that reported in normal NHPs and ET patients, and/or distinct clusters of SPN activity in our patients. To this end, we carefully identified single-unit activity from the striatum (putamen) of both PD and dystonic patients undergoing GPi-DBS surgeries and compared their discharge rates and patterns

## Results

### Database and spike sorting results

In this study, 93 microelectrode trajectories (48 in PD and 45 in dystonia) were used, yielding a total of 934 and 718 microelectrode recording (MER) segments within the striatum of patients suffering from PD and dystonia, respectively. Applying spike sorting on these MER segments identified 5336 units (3020 in PD and 2316 in dystonia). The MER segments and the sorted units from patients suffering from non-genetic (N = 27) and genetic (N = 18) dystonia were pooled, since no difference was detected between them. All surgeries were carried out while the patients were fully awake (no sedation or anesthesia) and the PD patients were off dopaminergic medication (overnight washout > 12 hours). The DBS target was the ventro-posterior-lateral portion of the GPi for all patients, and trajectory angles were only slightly modified according to patient’s anatomy. All patients provided their written informed consent and the study was approved by the Institutional Review Board of Hadassah Hospital in accordance with the Helsinki Declaration (reference code: 0168-10-HMO).

Fig.1A shows examples of the striatal spiking activity recordings (left and middle). The right plots display the superimposed waveforms of the extracellular action potentials of the sorted unit. The units are ordered as a function of their isolation score (Iso.Sc) (49). All recorded units were assigned to 10 bins based on their Iso.Sc and the median and mean discharge rates were calculated for each bin. A significant negative correlation between neuronal discharge rate and Iso.Sc was found in the striatum of the PD (Pearson’s r = −0.87 and −0.89 for median and mean discharge rate, respectively, P < 0.001, Fig.1B) and dystonic (Pearson’s r = −0.89 and −0.85 for median and mean discharge rate, respectively, P < 0.001, Fig.1B) patients, indicating that the units with the lowest Iso.Sc had the highest discharge rates.

**Fig.1.**
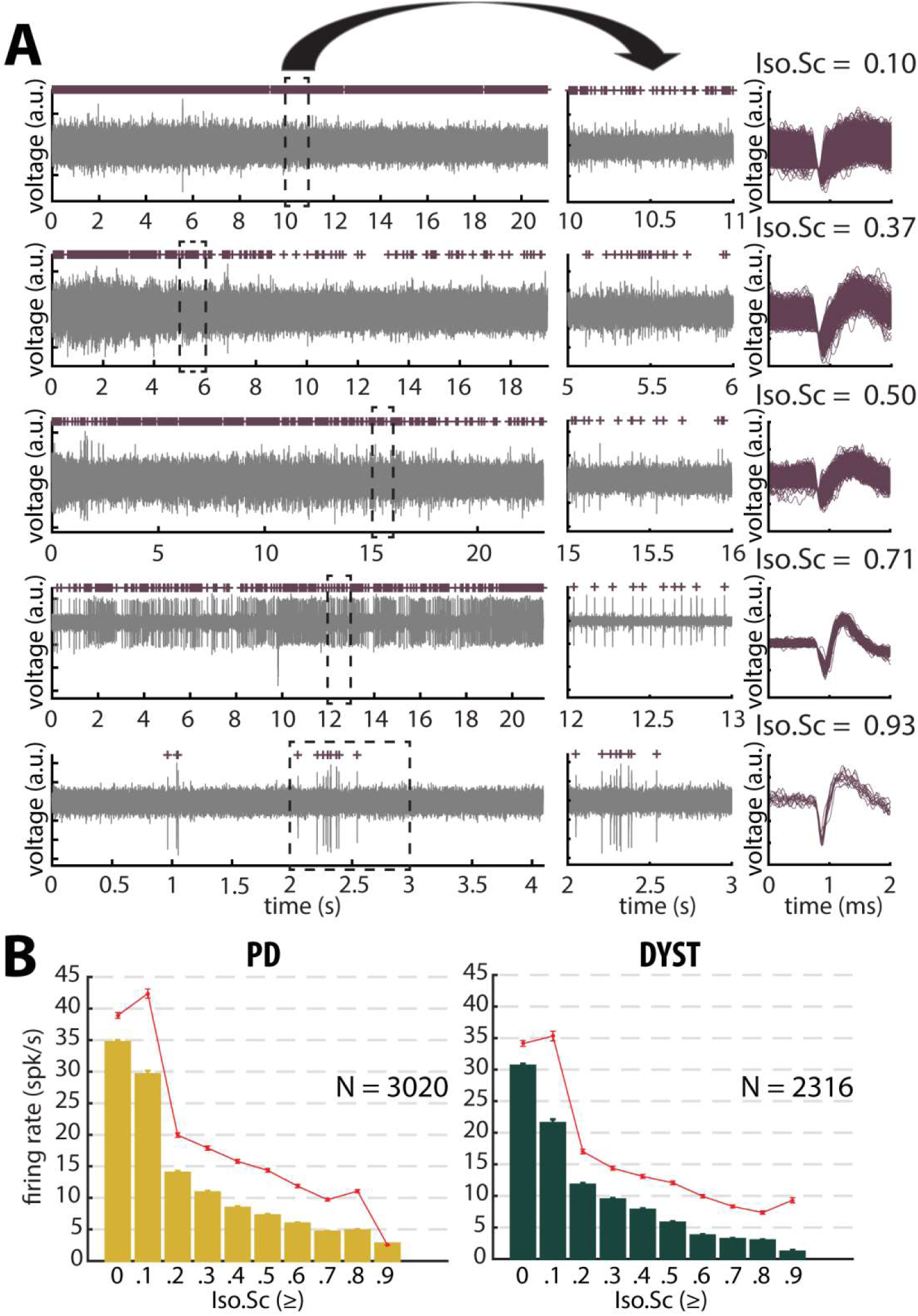
Striatal firing rate of parkinsonian (PD) and dystonic (DYST) patients decreases as isolation quality of the units increases. **(A)** Examples of full-length **(left panels)** and 1s (**middle panels)** striatal spiking recordings. Above the spiking activity is the digital display of the detections (spike train) of the sorted unit from the recording. Spike waveforms are superimposed **(right panels)**. Isolation score (Iso.Sc) indicates the isolation quality of the sorted unit. **(B)** Evolution of the firing rate as a function of the Iso.Sc. Each bar indicates the median firing rate of the sorted units with Iso.Sc greater than or equal to a certain value (i.e., the bin labeled 0.8 contains all the units with Iso.Sc ≥ 0.8). Error bars represent MADs (i.e., median absolute deviations). Means ± SEMs (i.e., standard errors of the mean) are shown in red. N is the number of sorted units.

### No dramatic increase in the firing rate of the striatal neurons of PD and dystonic patients

The median firing rate of all the sorted striatal units was relatively high (∼30-40Hz) in both diseases (Fig.2, first column) and consistent (at least for PD) with the values reported by Singh et al. (39). Similarly, we found that the firing rate of the dystonic patients was significantly lower than the PD firing rate (Mann-Whitney U-test, p < 0.001, Fig.S1, first column). Nevertheless, this significant difference disappeared and the striatal firing rate decreased dramatically when only comparing the firing rate of the well-isolated units (Iso.SC ≥ 0.6) (Fig.2 and Fig.S1, first column). There was therefore no dramatic or specific change in the striatal firing rate of PD and dystonic patients.

**Fig.2.**
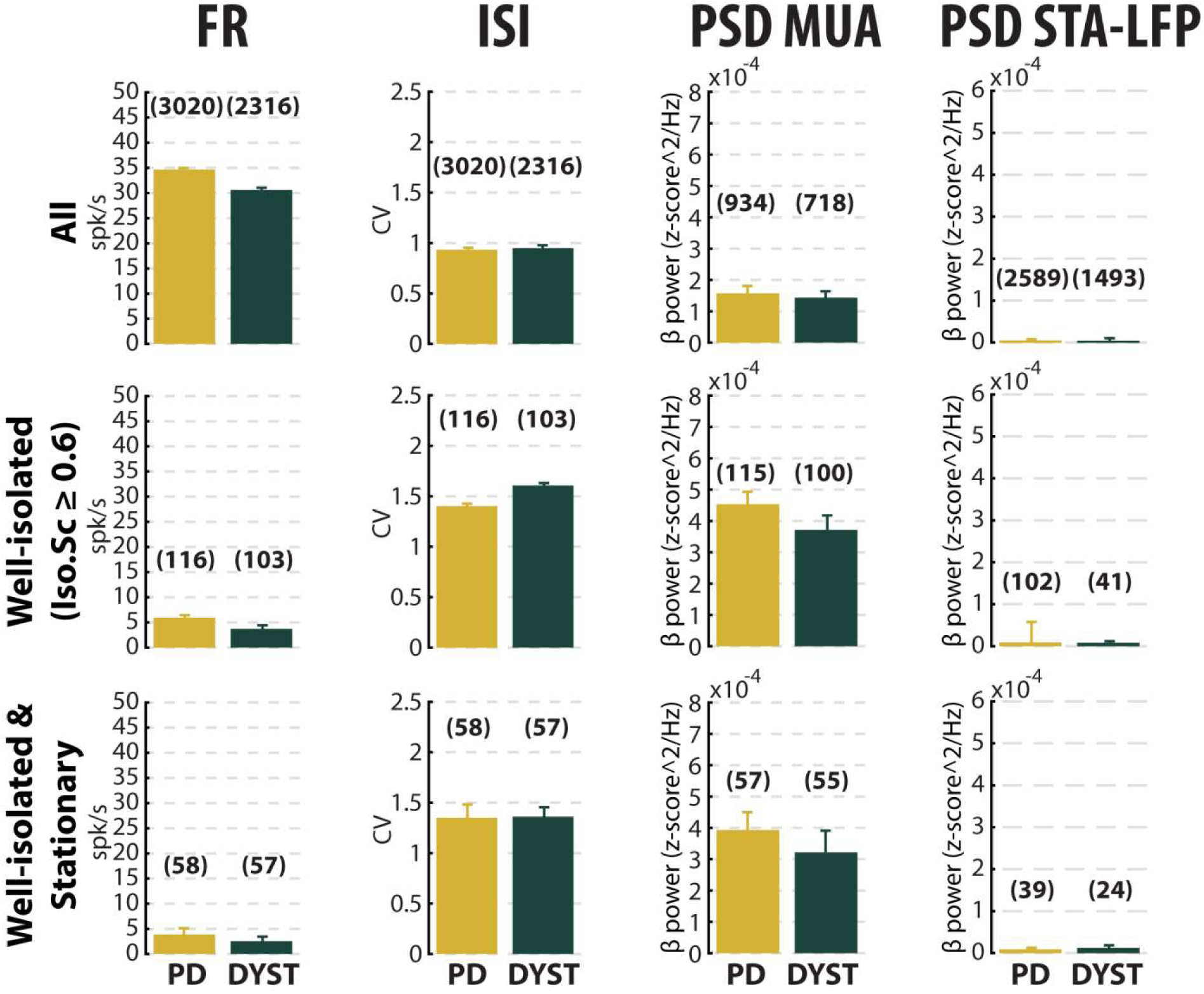
No dramatic or specific change in the features (median ± MAD values) of striatal neuronal activity in parkinsonian (PD) and dystonic (DYST) patients. Comparison of the firing rate **(first column)**, the coefficient of variation (CV) of the inter-spike interval (ISI) **(second column)**, the β power of the multi-unit activity (MUA or spiking activity) **(third column)** and the β power of the spike-triggered average (STA) of the local field potential (LFP) **(fourth column)** when considering all **(upper panels)**, only the well-isolated **(middle panels)** and only the well-isolated stationary **(lower panels)** units. Each bar indicates the median value and error bars represent MADs. Same y-axis scales as in Fig.S1. Numbers in parentheses indicate the sizes of the samples.

Inspection of the spike train of the well-isolated units revealed that although the units had been graded as high-isolation score units, their spike train could be non-stationary (Fig.S2A). To assess the stationary of the spike trains, we examined the temporal (linear) evolution of their firing rate and spike amplitude (see Materials and Methods). Fig.S2A shows 3 representative examples of well-isolated non-stationary units (left panels) and 3 representative examples of well-isolated stationary units (right panels) from the striatum of PD patients. Here, we used the slopes of the linear regression lines (reflecting the temporal evolution of the firing rate and spike amplitude of the well-isolated units) to define stationary units (Fig.S2B and C). For both PD and dystonia, we found that the firing rate of the stationary units was significantly lower than the firing rate of the most non-stationary units (Mann-Whitney U-test, p < 0.01 and 0.001 for PD and dystonic patients, respectively, Fig.S2D). Thus, inclusion of non-stationary units could have erroneously inflated the striatal firing rate in PD and dystonic patients. Removing the non-stationary units reduced the striatal firing rate of the well–isolated units in the striatum of both PD and dystonic patients (Fig. 2 and Fig.S1 first column). Moreover, the striatal firing rate remained similar between both diseases after this maneuver.

Equally important, we found that mean values of the firing rate (Fig.S1, first column) were systematically higher than the median values (Fig.2, first column), suggesting that whatever the quality of the isolation and the stationarity of the striatal units, the distributions of the firing rate were not normal and were skewed to the right. Fig.S3 depicts the distributions of the firing rate of all (Fig.S3A) and only the well-isolated (Fig.S3B) units. For both PD and dystonia, and for all the levels of spike isolation, the distributions of the firing rate of the SPNs were not normally distributed, but were rather strongly skewed to the right (Fig.S3B). We therefore used the median (Fig.2) rather than the mean (Fig.S1) to represent the central moment of the distribution of the discharge properties (including the discharge rate) of the SPNs. The median ± MAD (median absolute deviation) of the discharge rate was 3.85 ± 1.18Hz and 2.55 ± 0.81Hz for the well-isolated stationary SPNs in PD and dystonia respectively - i.e. in the same range as reported for the controls in animal studies and with no significant difference between PD and dystonia.

### No evidence for bursty patterns in the striatal spiking activity of PD and dystonic patients

To characterize the pattern of the spike trains (i.e., irregular, periodic or bursty) and compare them between PD and dystonic patients, we examined the time interval histograms (TIHs) of the inter-spike intervals (ISIs). Although weak (Fig.2, second column), we found a significant difference in the coefficient of variation (CV) of the ISIs of all the sorted striatal units between PD and dystonic patients (Mann-Whitney U-test, p < 0.001, Fig.S1, second column). Nevertheless, as for the striatal firing rate, this difference vanished when only considering the well-isolated or well-isolated stationary units (Fig.2 and Fig.S1, second column). Again, the mean values of the CV of the ISIs (Fig.S1 second column) were systematically higher than the median values (Fig.2, second column), indicating skewed distributions. The distributions of TIHs of the ISIs of the well-isolated stationary units matched the Poisson temporal distribution of spikes (Fig.S4A). Accordingly, the mean autocorrelograms did not reveal any periodic or bursty firing patterns in the striatal well-isolated stationary units (Fig.S4B). Similarly, the prevalence of bursts was relatively low (Fig.S4C) and did not significantly differ between PD and dystonic patients (Mann-Whitney U-test, p > 0.05, Fig.S4D).

### Absence of oscillatory spiking activity in the striatum of PD and dystonic patients

To overcome possible confounding effects of the low discharge rate and spatial under-sampling of the striatal SPNs, we investigated multi-unit oscillatory activity rather than single-unit oscillatory activity. Striatal multi-unit activity (MUA) reflects the spiking activity of an ensemble of striatal neurons. Theoretical studies have shown that neural oscillations can emerge at the population level in networks of neurons exhibiting an irregular (i.e., non-oscillatory) discharge pattern and a low firing rate (50, 51).

Comparison of the power spectral densities (PSDs) of the spiking activities recorded in the striatum of the PD and dystonic patients did not reveal any oscillatory phenomena between 3 and 75Hz, including the β (13-30Hz) band (Fig.S5A and B). Accordingly, no significant difference in the β power of the striatal spiking activities (whatever the quality of the isolation and the stationary of the striatal units) was observed between PD and dystonic patients (Mann-Whitney U-test, p > 0.05, Fig.2, and Fig.S1 third column), Moreover, we found no significant increases of the β (13-30Hz) power in the spiking activities recorded in the vicinity of striatal well-isolated stationary units compared to the linearly interpolated baseline β power (Wilcoxon signed rank test, p > 0.05, Fig.S5C), supporting the view of no oscillatory spiking activity in the striatum of PD and dystonic patients.

### No locking between spike and β LFP oscillations in PD and dystonic patients

The mean PSD of the mono-polar LFPs recorded in the striatum of PD and dystonic patients (Fig.S6A) exhibited (ignoring 50Hz artifacts) two and three distinct peaks, respectively (at 12 and 22Hz for PD patients and 9, 23 and 31Hz for dystonic patients, Fig.S6A, insets). Therefore, we further calculated the spike-triggered averages of the LFPs (STAs LFPs). The β power in the LFPs recorded around the time of the spikes of all the striatal sortedunits was significantly higher in PD patients than in dystonic patients (Mann-Whitney U-test, p > 0.0 1, Fig.2 and Fig.S1, fourth column). However, this difference disappeared when using the spikes of the well-isolated (non-stationary and stationary or only stationary) units (Fig.2 and Fig.S1, fourth column). Thus, the spiking activity of well-isolated stationary units recorded in the striatum of both PD and dystonic patients failed to lock with 13-30Hz LFP oscillations (Fig. S6B).

### Cluster analysis of striatal discharge

To further test whether two distinct SPN subpopulations could be observed in the current study, we performed 2D k-means cluster analysis with k=2, using the discharge rate and the CV of the ISIs of each unit as input parameters (Fig. 3). Using k=2 as predefined numbers of clusters systematically enforces the separation into two distinct SPN subpopulations; however calculation of the silhouette values revealed that the two clusters were not well-separated, whatever the quality of the isolation and the stationarity of the striatal units (Fig. 3, insets). Equally important, it was clear that the tendency to burst [CV of the ISIs >1 (52)] decreased for units with a high discharge rate (Fig. 3). Similar 2D or higher-order (3 or 4D) k-means cluster analyses (using the discharge rate, CV of the ISIs, MUA β power or STA-LFP β power as input parameters) also failed to reveal two well-separated clusters.

**Fig.3.**
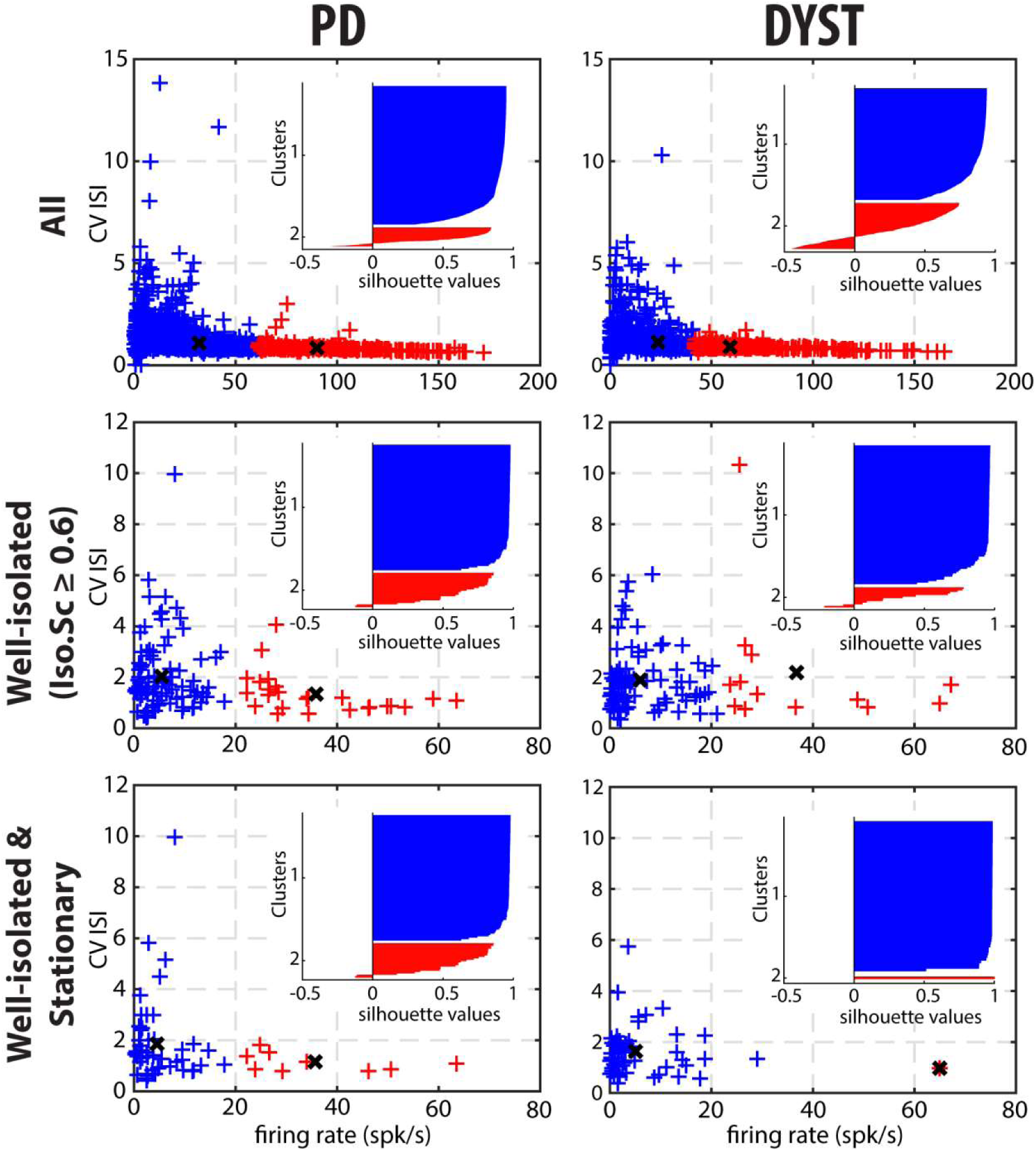
Cluster analysis using discharge properties does not reveal well-separated subpopulations of striatal units in parkinsonian (PD) and dystonic (DYST) patients. 2D k-means cluster analysis with k=2, using the firing rate and the CV of the ISIs of each unit as input parameters. Analysis was performed when considering all **(upper panels)**, only the well-isolated **(middle panels)** and only the well-isolated stationary **(lower panels)** units. Markers (x) represent cluster centroids. Inset: silhouette values (range between −1 and 1) were calculated for each clustering to assess how well-separated the two resulting clusters were.

## Discussion

There is no consensus as to the impact of dopamine depletion on the discharge rate and pattern of the SPNs. (25, 38–40, 43, 44, 46). Recordings of the activity of the SPNs are inherently difficult to perform given their very low firing rate and small size, leading to frequent loss and damage of these units (53). These difficulties can distort the reported population averages of SPNs. For these reasons, we used an automated data driven approach. A total of 934 and 718 MER segments from the striatum of PD and dystonic patients were analyzed. The striato-pallidal border was automatically detected using a machine learning algorithm (54). Spike detection was performed offline with fully automatic quantification of the isolation quality and stationarity of the identified units. Unlike previous reports (39), we found no drastic modulation in the SPN discharge rate and pattern (compared to values reported for normal/control animals or ET patients) in PD and dystonic patients.

### Low discharge rate and similar levels of striatal activity in PD and dystonia

We found that the firing rate of the SPNs of both the PD and dystonic patients was extremely sensitive to the quality of the isolation of the units. Specifically, we found a negative correlation between striatal firing rate and isolation score (Fig.1A and B). The negative correlation between the discharge rate of the detected units and their isolation quality revealed that - unlike in STN (55) - the spike detection and sorting algorithm used in this study and/or the physical properties of the striatal neurons tended to erroneously classify noise events as spikes (false positives) rather than missing real spikes (false negatives). Removing the most non-stationary units also reduced the striatal firing rate of SPNs in both PD and dystonic patients (Fig.2, first column), thus further indicating that the inclusion of non-stationary (“injured”) units can also erroneously increase the striatal firing rate in PD and dystonic patients. This automatic approach to spike detection, sorting and quality assessment revealed that the median firing rate of the striatal well-isolated stationary units was 3.85 and 2.55Hz in PD and dystonic patients, respectively (Fig.2, first column).

Our results are inconsistent with a recent study by Singh et al. (39) where the SPN discharge rate in PD (30.2 ± 1.2Hz) and dystonic (9.3 ± 0.6Hz) patients increased by ∼15- and 5-fold compared to the low striatal discharge rate (2.1 ± 0.1Hz) found in patients suffering from ET (i.e., a disorder without any known BG malfunction). In the current study, we did not have a control condition, nor did we record the striatal activity from ET patients. Therefore, we cannot rule out the possibility of an increase in the firing rate in the striatum of both PD and dystonic patients compared to human controls. Moreover, we did not differentiate between SPNs and striatal interneurons (including the TANs). About 95% of the striatal neurons are SPNs and only ∼2% are cholinergic interneurons (presumed TANs) [see e.g., (56)]. We therefore considered our sorted striatal spiking activity to reflect SPN spiking activity and neglected the small fraction of TANs probably included in our sample. In any case, we did not find any significant difference between PD and dystonic patients that might reflect a distinct level of striatal hyperactivity in PD (Figs 2 and S1, first column). These different results may be due to: (i) a difference between automatic and manual striato-pallidal border demarcation, (ii) the use of algorithms for the quantification of the isolation quality and stationarity of the units, and (iii) different recording techniques (microelectrode type and step size) and/or spike detection methods.

### Lack of SPN bursty and oscillatory pattern in the SPN spiking activity of PD and dystonic patients

The emergence of bursty and periodic oscillatory patterns after striatal dopamine depletion has been observed in other BG nuclei than the striatum (25, 26, 34, 57, 58). Previous studies by our research group reported the emergence of oscillatory spiking activity in the TANs (presumably the striatal cholinergic interneurons) of the MPTP-treated monkey, thus indicating that abnormal oscillatory activity following striatal dopamine depletion did not spare the striatum (25, 59). However, we found no significant parkinsonism-related oscillations in SPN spiking activity (25, 34). Here, in line with this finding, we did not observe the emergence of bursting (Figs 2 and S1, second column and Fig.S4) or oscillatory (Figs 2 and S1, third column and Fig.S5) spiking activity of the striatal units sorted in the PD and dystonic patients.

In contrast to our results, Singh and collaborators reported large increases in the bursting activity of the SPNs in PD and dystonic patients (39) and MPTP-treated monkeys (40). Levodopa-induced increases or decreases of the SPN firing rate may be the result of D1 or D2 receptor activation, respectively (60–62). Using the recognized responses to dopaminergic stimulation, Singh et al. (2015) (40) showed that most bursty SPNs in the parkinsonian NHP exhibited a D1 receptor response to levodopa, thus suggesting that the emergence of an abnormal firing pattern predominantly affected the D1 SPNs (i.e., the SPNs of the direct pathway). However, when using a single-cell juxtacellular recording–labeling technique in the 6OHDA rodent, Sharott et al. showed that in addition to their excessive firing rate, D2 SPNs (that primarily composed the indirect pathway) displayed aberrant phase-locked burst firing to cortical β oscillations (44). In that study, one might assume that D2 SPNs, rather than D1 SPNs, would exhibit an abnormal firing rate and pattern in the dopamine-depleted striatum. Therefore, technical advances and further studies in PD patients and animal models should be carried out to reach a consensus.

As reported above (25, 29–32, 34, 63), abnormal oscillations of LFP were recorded in the striatum of both PD and dystonic patients (Fig.S6A). However, we found no significant phase-locking between striatal spiking activity and mono-polar LFP β oscillations (Figs 2 and S1 fourth column and Fig.S6B). These results support the view that β oscillatory spiking activity is not materialized in the SPNs and that the BG oscillatory activity observed in both PD and dystonic patients does not resonate through the striatum. BG LFP represents sub-threshold (e.g., synaptic input) activity at best (64, 65). Recent studies have demonstrated that monopolar and bipolar BG LFPs may be contaminated by the volume conductance of cortical electroencephalogram (EEG) activity (66, 67). Therefore, it is likely that BG LFPs, including the LFPs recorded in the striatum, do not accurately reflect local cellular activity and should be at best interpreted with caution. Exaggerated striatal LFP oscillations in PD and dystonia cannot therefore be regarded as direct evidence for the presence of SPN spiking oscillatory activity.

### Is the imbalance in the activity of SPN subpopulations evident in PD and dystonia?

Obviously, striatal activity must be affected in PD and dystonic patients. One claim suggests that hypoactivity of the direct pathway (originating from D1 SPNs) and hyperactivity of the indirect pathway (originating from D2 SPNs) lead to excessive GPi/SNr inhibitory inputs to the thalamus in PD (27) and vice versa in dystonia (28). These aberrant GPi/SNr inhibitory inputs to the thalamus lead to the release of abnormal output commands and result in the emergence of the clinical symptoms. Unlike in rodent studies [see e.g., (44)], extracellular recording methods cannot discriminate between the spiking activity of the striatal D1 and D2 SPNs. However, if striatal dopamine depletion drastically enhances the differences (in discharge rate and pattern) between the two distinct SPN subpopulations [but see (46)], one would expect to see a bimodal distribution of SPN discharge properties after dopamine depletion. Instead, visual inspection of the distributions of the discharge rate (Fig.S3) and the CVs of ISIs (Fig.S4A) revealed long-tailed unimodal distributions in both diseases. Moreover, the cluster analysis using SPN discharge properties failed to identify two well-separated SPN subpopulations.

In conclusion, our current results in patients extend our previous study in the MPTP NHP model of PD (25) and studies in the 6-OHDA rodent model [e.g.(46, 48)] and demonstrate that abnormal activity along both the direct and indirect pathways of the BG network is not caused by dramatic changes in SPN spiking activity. Therefore, we propose that there is an abnormal recruitment (e.g., by behavioral events) of D1 and D2 SPNs (which are mostly silent at rest in the healthy condition) in pathological conditions that result in an aberrant net balance of striatal outputs (48). Moreover, it is likely that these moderate changes in SPN discharge are amplified by BG downstream structures, thus leading to the clinical symptoms of PD and possibly of dystonia.

## Materials and Methods

### Patients and surgery

Patients with PD and dystonia were recruited from the movement disorders clinics at the Hadassah Medical Center in Jerusalem. All patients were scheduled to undergo implantation of DBS electrodes into the GPi and underwent MR imaging, and evaluation for motor and non-motor impairments within the 30 days prior to surgery. Data were collected from 16 PD and 13 dystonic (non-genetic and genetic dystonia) patients. Patient demographic information appears in table S1. Note that our DBS recordings in ET patients start 10mm above the thalamic target and therefore do not include striatal recordings. All patients met the criteria for DBS and signed a written informed consent for surgery that involved microelectrode recording. This study was authorized and approved by the Institutional Review Board of Hadassah Hospital in accordance with the Helsinki Declaration (reference code: 0168-10-HMO)

Surgery was performed using a CRW stereotactic frame (Radionics, Burlington, MA, USA). BG target coordinates were chosen as a composite of the indirect anterior commissure-posterior commissure (AC-PC) atlas-based location and direct (1.5 or 3Tesla) T2 magnetic resonance imaging, using Framelink 4 or 5 software (Medtronic, Minneapolis, USA). All recordings used in this study were made while the patients were fully awake (no sedation or anesthesia) and PD patients were off dopaminergic medication (overnight washout > 12 hours).

### Data acquisition

The data were acquired using two systems: MicroGuide [prior to 2015, previously described (55) and Neuro Omega (from 2015, previously described (54)]

### Microelectrode recordings

For every recording session, a microelectrode recording (MER) exploration using one or two microelectrode trajectories (2 mm apart) was made starting at 15 mm above the pre-operative T2 MRI image-based calculated target. Our trajectories followed a double-oblique approach (50 to 80° from the axial AC-PC plane and 0 to 10° degrees from the mid-sagittal plane) through the striatum and the GPe and towards the ventral border of the posterior-lateral portion of the GPi target. The target coordinates were in the range of the approximate anatomic coordinates for the GPi: Lateral (*X*) = 19–22 mm from the midline or ∼18 mm from the third ventricle wall; Anterior (*Y*) = 1–3.5 mm from the mid-commissural point (MCP), and vertical (*Z*) = −1 to −4 below the AC-PC plane.

The “central” electrode was directed at the ventral border of the posterior-lateral portion of the GPi and the “anterior” electrode was located 2mm anterior/ventral to the central electrode in the parasagittal plane. Some of these recordings were made by a single microelectrode trajectory (instead of two) to accommodate cortical anatomy under the burr hole and brain blood vessels (68).

MER segments were regularly sampled in space. For all our GPi-DBS surgeries, the step size between two MER segments ranged from 100 to 200µm and was controlled by the neurophysiologist to achieve optimal identification of the pallidal borders. At each step, MER segments were recorded from 4 to 140s (after a 2-s signal stabilization period).

A total of 93 microelectrode trajectories aiming at the GPi were analyzed (48 in PD and 45 in dystonia), yielding a total of 934 and 718 MER segments within the striatum of patients suffering from PD and dystonia, respectively. Striatum-GPe borders were automatically detected using a machine learning software (54).

### Offline spike sorting and assessment of isolation quality and stationarity of identified/sorted units

Single-unit striatal activity was assessed by sorting spike trains from every MER segment recorded within the striatum via an automatic offline spike detection and sorting method (Offline Sorter v4.4.2.0, Plexon Inc, Dallas, Texas). The threshold-crossing method for spike detection (a negative voltage threshold trigger systematically set at 3SDs from the mean of the peak height histograms), principal component analysis for spike feature extraction and k-means cluster method (K-Means Scan with number of clusters between 1 and 5) were used. Quantification of the isolation quality of the spike detection was obtained by calculating the isolation score (Iso.Sc) (49). Iso.Sc ranges from 0 (i.e. highly noisy) to 1 (i.e. perfect isolation).

In order to assess the stationarity of the firing rate and the spike amplitude of the well-isolated units (Iso.Sc ≥ 0.6), the firing rate and the average spike amplitude of each well-isolated unit were Z-normalized over 10 bins (from the first to the last spike of the spike train). Then, the slope value of the linear regression line for the firing rate and the average spike amplitude were calculated and expressed as the z-score/bin. Units with slope values of the linear regression lin. e for firing rate or average spike amplitude greater than or equal to the 70th percentile were defined as non-stationary units.

### Discharge pattern assessment of spike train

For each spike train of the well-isolated stationary unit, we calculated the inter-spike intervals (ISI) and generated the ISI histograms of the well-isolated stationary units. In parallel, we also computed the autocorrelograms of the spike train of the well-isolated stationary units, calculated for ±500 ms offset with 10ms-bins. For each autocorrelogram, values were normalized so that autocorrelation values ranges from 0 to 1.

For burst detection, we applied the Poisson surprise method with the surprise maximization (SM) search algorithm (69) to each spike train of the well-isolated stationary units, using the following parameters: minimal burst length = 3 spikes; threshold surprise value (S) = 10 (i.e., only bursts having S>10, corresponding to an occurrence rate of ∼ 0.01 bursts/1000 spikes in a random spike train, were considered to be of interest); burst ISI limit = mean (ISI)/ 2; add limit = 150% of burst ISI limit and inclusion criteria (IC) = 5.

For each spike train of the well-isolated stationary units, the frequency (number of bursts/s) and mean duration (s) of the burst episodes were calculated over their entire recording span. These two metrics were used to determine the burst prevalence for each unit which was defined as the: burst frequency * mean burst duration. For each unit, the burst prevalence (range: 0-1) represents the probability that the discharge pattern is bursty.

### Power spectral density

For the power spectral density (PSD) calculations, the band-pass filtered spiking signal was Z-score normalized to obtain an unbiased estimate (by the electrode impedance, the A/D characteristics of the recording system, or the amplitude of the recorded neuronal activity) of the oscillatory activity (37). The Z-normalized signal was rectified by the “absolute” operator (25, 34, 37, 55, 70). The rectified signal follows the envelope of the MUA and therefore enables the detection of burst frequencies below the range of the online band-pass filter. Since the LFP frequency domain was filtered out, the resulting PSD only represented the oscillatory features of the spiking activity.

The PSD of each multi-unit site was calculated using Welch’s method with a 1-s Hamming window (50% overlap) and a spectral resolution of 1Hz (nfft = 44000 or 48000, sampling frequency = 44 or 48kHz depending on the acquisition system). To evaluate the β power, the baseline values in the 13-30Hz range of each PSD were linearly interpolated [based on the two closest points that flanked the 13-30Hz band, namely the values at 12 and 31Hz (spectral resolution of 1Hz, see above)] and averaged. Then, the β power (i.e., the mean of the observed values between 13 and 30Hz) was compared to the interpolated β power (baseline).

Similarly, we also calculated the PSD of the striatal monopolar (0-300Hz) LFP. To do so, the PSD of each Z-normalized LFP was calculated using Welch’s method (see above for the parameters; but nfft = 1375, sampling frequency = 1.375kHz) and without prior rectification by the absolute operator

### Spike-triggered average of the LFP

To investigate the spike-LFP relationship in the temporal domain, we also calculated the spike-triggered average (STA) of the LFP (25, 71) for well-isolated stationary units. In doing so, the LFP was recorded in the vicinity of well-isolated stationary units (i.e., spiking activity and LFP were recorded on the same electrode). Each Z-normalized LFP was offline band-pass filtered from 4 to 12Hz or from 13 to 30Hz (4-pole Butterworth filter, filtfilt Matlab function). For comparison, STAs-LFP were also calculated after randomly shifting the timestamp of each spike of the spike train [i.e., random time (comprised between 0 and 1s) was added to the timestamp of each spike of the spike train] in order to abolish any relationship between LFP and spiking activity (Shift predictor). The PSD of the STA-LFP was calculated as for the LFP.

### Software and Statistics

All the data and statistical analyses were carried out using custom-made MATLAB R2016a routines (Mathworks, Natick, MA, USA). Mann-Whitney U-test and Wilcoxon signed rank tests were used for statistical comparisons of paired and unpaired samples, respectively. The criterion for statistical significance was set at P < 0.05 for all statistical tests

### Data availability

The data are available from the corresponding author upon request.

## Data Availability

The data are available from the corresponding author upon request.

## Footnotes

Author contributions: D.V., Z.I, T.B., H.B. and M.D. designed the research; D.V., M.D. performed the research; Z.I. carried out the surgery; D.V. and M.D. analyzed the data; D.V., H.B. and M.D. wrote the paper. All authors read and approved the final version.

The authors declare no conflict of interest.

* To whom correspondence should be addressed.

\# Present address: Institute of Neurodegenerative Diseases - CNRS UMR 5293, University of Bordeaux, Centre Broca Nouvelle-Aquitaine - 3ème étage 146 rue Léo Saignat - CS 61292 - Case 28 33076 Bordeaux cedex, France

## Acknowledgments

This study was supported by the European Research Council (ERC), Rosetrees and Magnet grants to H.B. and The French National Research Agency (ANR) French National Center for Scientific Research (CNRS) to M.D. We thank Esther Singer for editing.

## Supplementary information

**Fig.S1.**
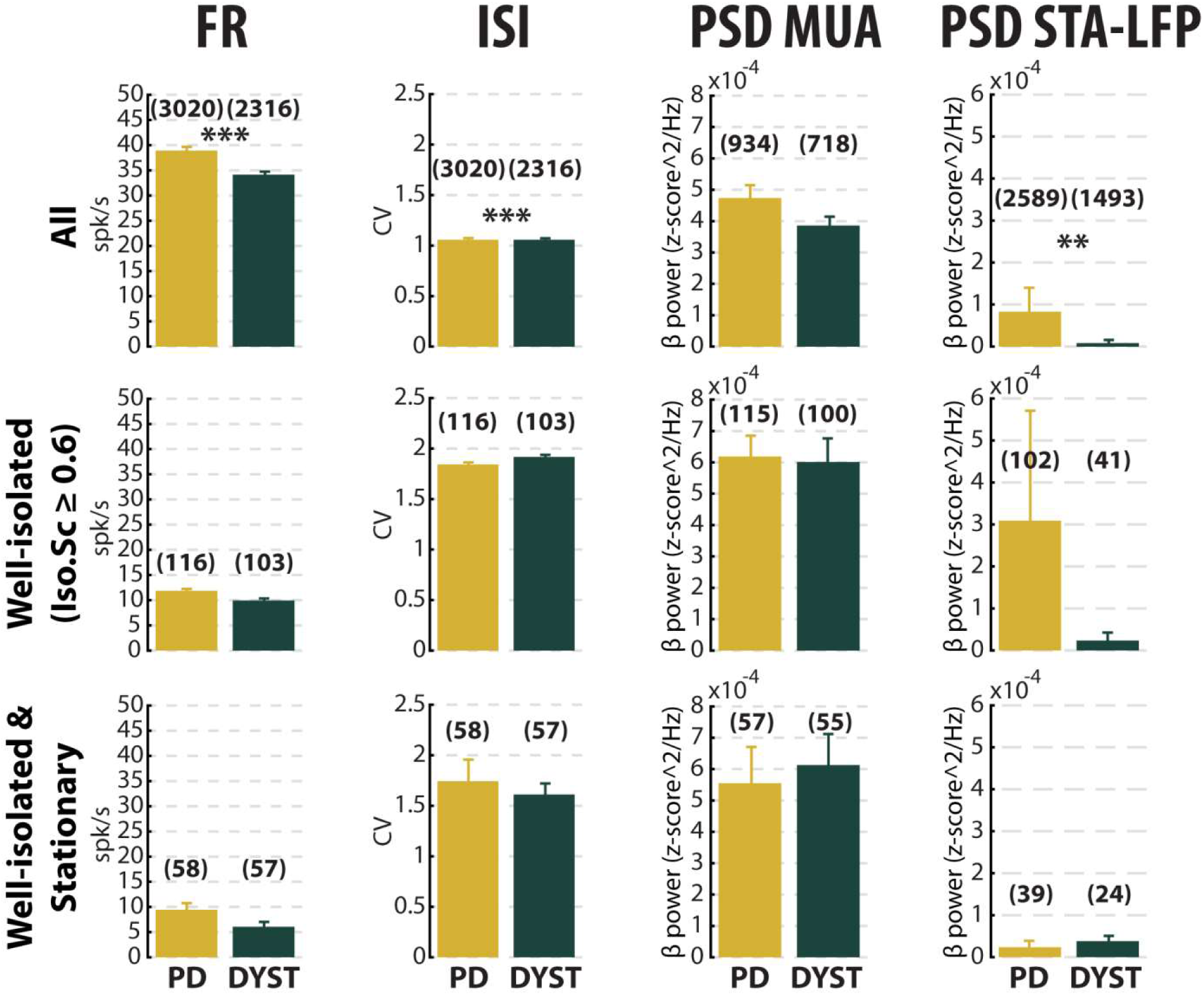
No dramatic or specific change in the features (mean ± SEM values) of striatal neuronal activity in parkinsonian (PD) and dystonic (DYST) patients. Same convention as Fig. 2, except that each bar indicates the mean value and error bars represent SEMs. Same y-axis scales as in Fig.2. **, *** indicate significant (p < 0.01 and 0.001, respectively) differences (Mann-Whitney U-test).

**Fig.S2.**
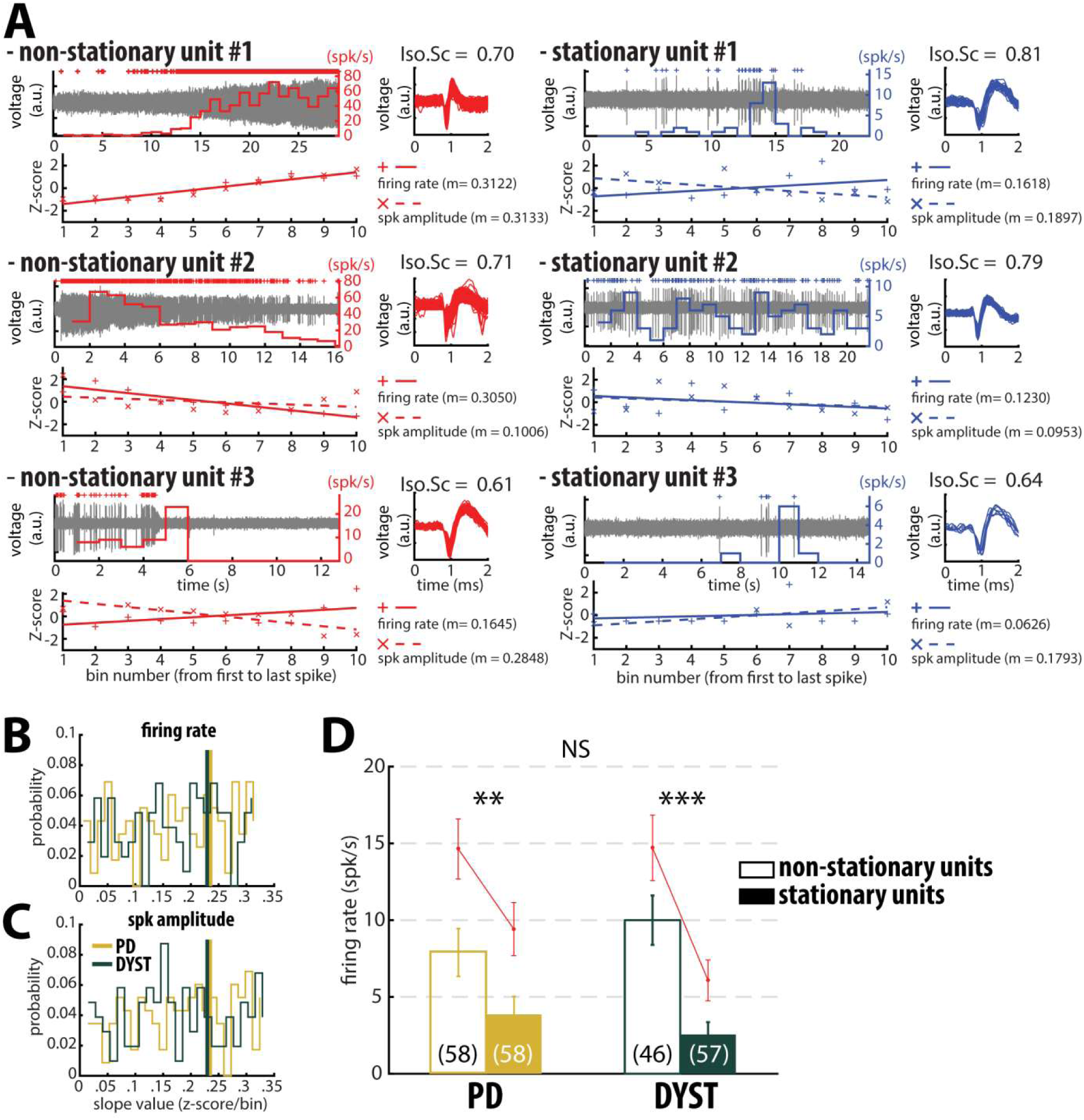
Inclusion of non-stationary units can erroneously increase the striatal firing rate in parkinsonian (PD) and dystonic (DYST) patients. **(A)** Examples of well-isolated (non-stationary and stationary) units recorded in the striatum of PD patients. Grey trace is the band-pass filtered signal and depicts the spiking activity. Above the spiking activity is the digital display of the detections (spike train) of the sorted unit. The firings rate of the sorted unit overlays the spiking activity. The spike waveforms extracted from the spiking activity are superimposed and displayed on the right. Iso.Sc indicates the isolation quality of the identified unit. Below the spiking activity panel is the assessment of the stationarity of the firing rate and the spike amplitude. The firing rate and average spike amplitude of the sorted unit were Z-normalized over 10 bins (from the first to the last spike of the spike train). Solid and dotted lines represent the linear regression lines (m) between the firing rate/spike amplitude of the sorted unit and bin number. Slope value (z-score/bin) of the linear regression line is used to assess stationarity. **(B-C)** Distributions of the slope values of the linear regression line for the firing rate and the spike amplitude of the units. Vertical lines indicate the 70th percentile for the two diseases (PD: 0.2355 and 0.2347 z-score/bin for firing rate and spike amplitude, respectively; dystonia: 0.2292 and 0.2294 z-score/bin for firing rate and spike amplitude, respectively). Units with slope values of the linear regression line for firing rate or spike amplitude greater than or equal to the 70th percentile were defined as non-stationary units. **(D)** Comparison of the firing rate of the non-stationary and stationary units. Each bar indicates the median firing rate of the sorted units. Error bars represent MADs. Means ± SEMs are shown in red. Numbers in parentheses indicate the numbers of non-stationary and stationary units. **, *** and NS indicate significant (p < 0.01 and 0.001) and non-significant differences, respectively (Mann-Whitney U-test).

**Fig.S3.**
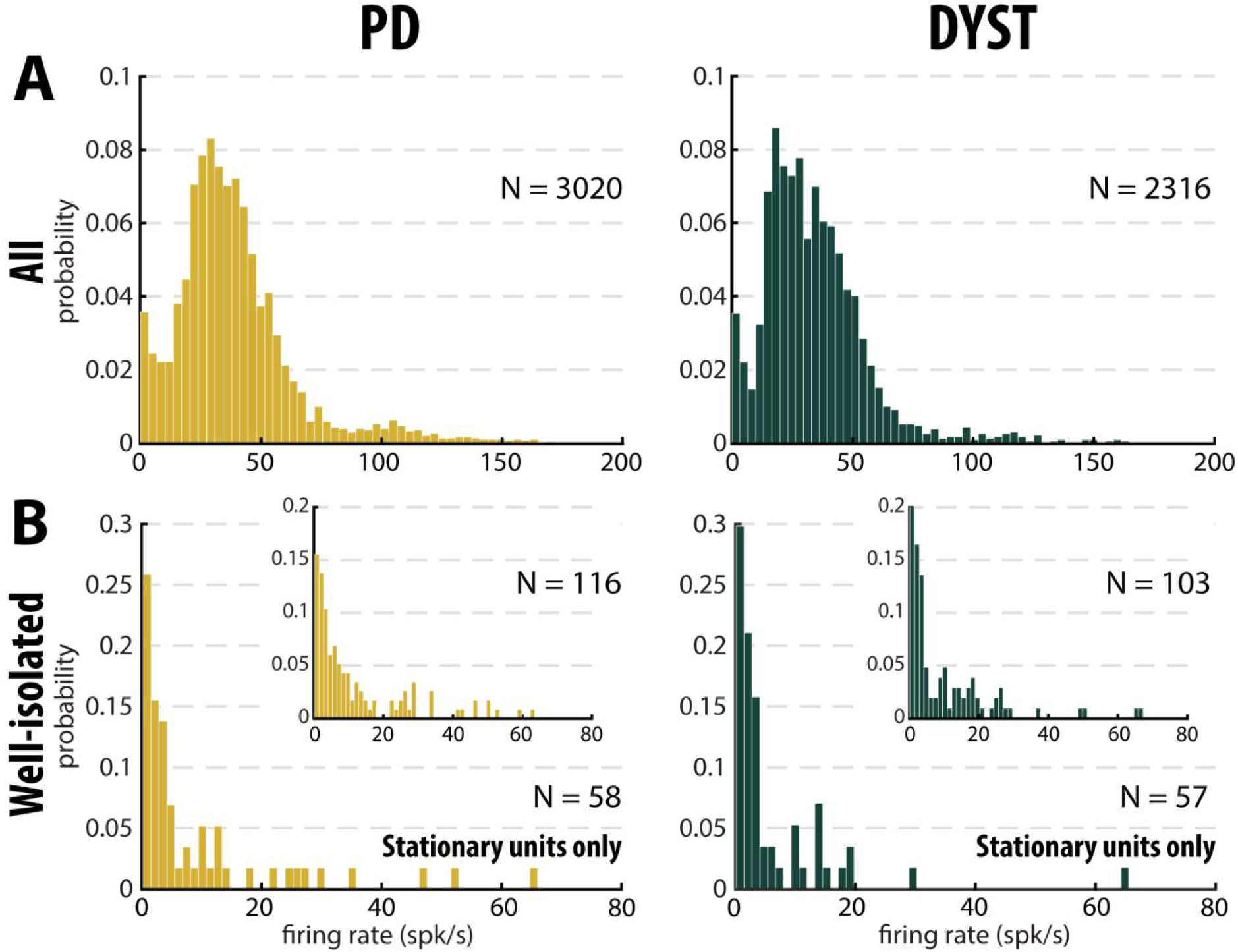
Distributions of the striatal firing rate are skewed to the right in parkinsonian (PD) and dystonic (DYST) patients. **(A)** Firing rate of all sorted units regardless of their isolation quality. **(B)** Firing rate of the well-isolated stationary units only. Skewness = 2.38 and 3.88 for PD and dystonic patients, respectively). Insets: Firing rate of the well-isolated units (non-stationary and stationary units pooled). Skewness = 1.74 and 2.36 for PD and dystonic patients, respectively. N is the number of sorted units

**Fig. S4.**
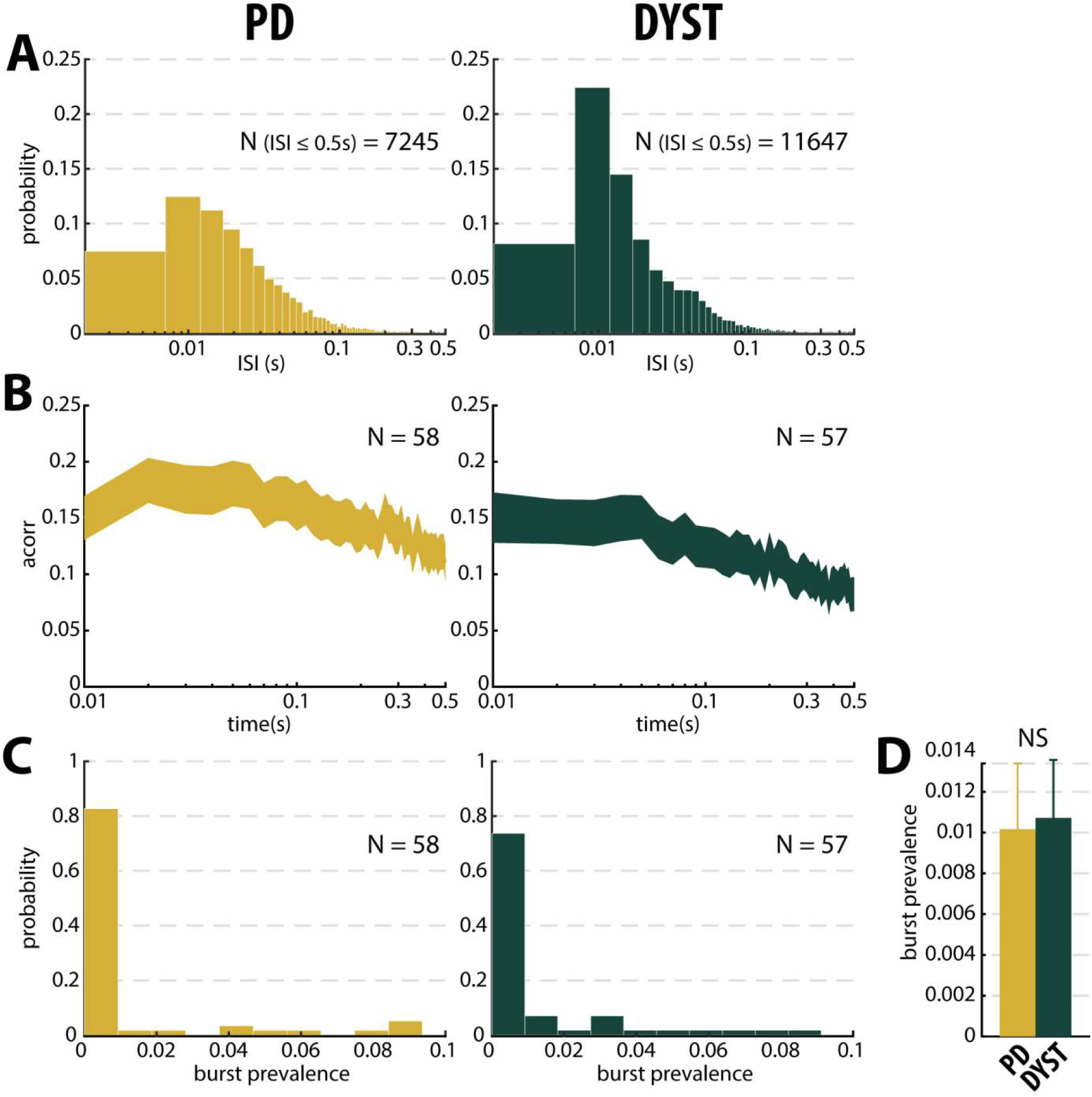
No evidence for burst patterns in the striatal spiking activity of parkinsonian (PD) and dystonic (DYST) patients. **(A)** Time interval histograms of the inter-spike intervals (ISI) of the well-isolated (Iso.Sc ≥ 0.6) stationary units. For better visualization ISIs > 0.5s were removed (191 and 292 for PD and DYST striatal units, respectively). Abscissas are in log scale. **(B)** Average (mean ± SEM) autocorrelograms of the well-isolated stationary units. For each autocorrelogram, values were normalized so that the autocorrelation values ranged from 0 to 1. Abscissas are in log scale. N is the number of well-isolated stationary units averaged. **(C)** Distributions of the values of burst prevalence for the spike train of the well-isolated stationary units. For each unit, episode prevalence represents the probability that the discharge pattern was bursty. **(D)** Mean values of burst prevalence for the well-isolated stationary units recorded in the striatum of parkinsonian (PD) and dystonic (DYST) patients. Error bars represent SEMs. N is the number of well-isolated stationary units averaged. NS: Non significant (Mann-Whitney U-test).

**Fig.S5.**
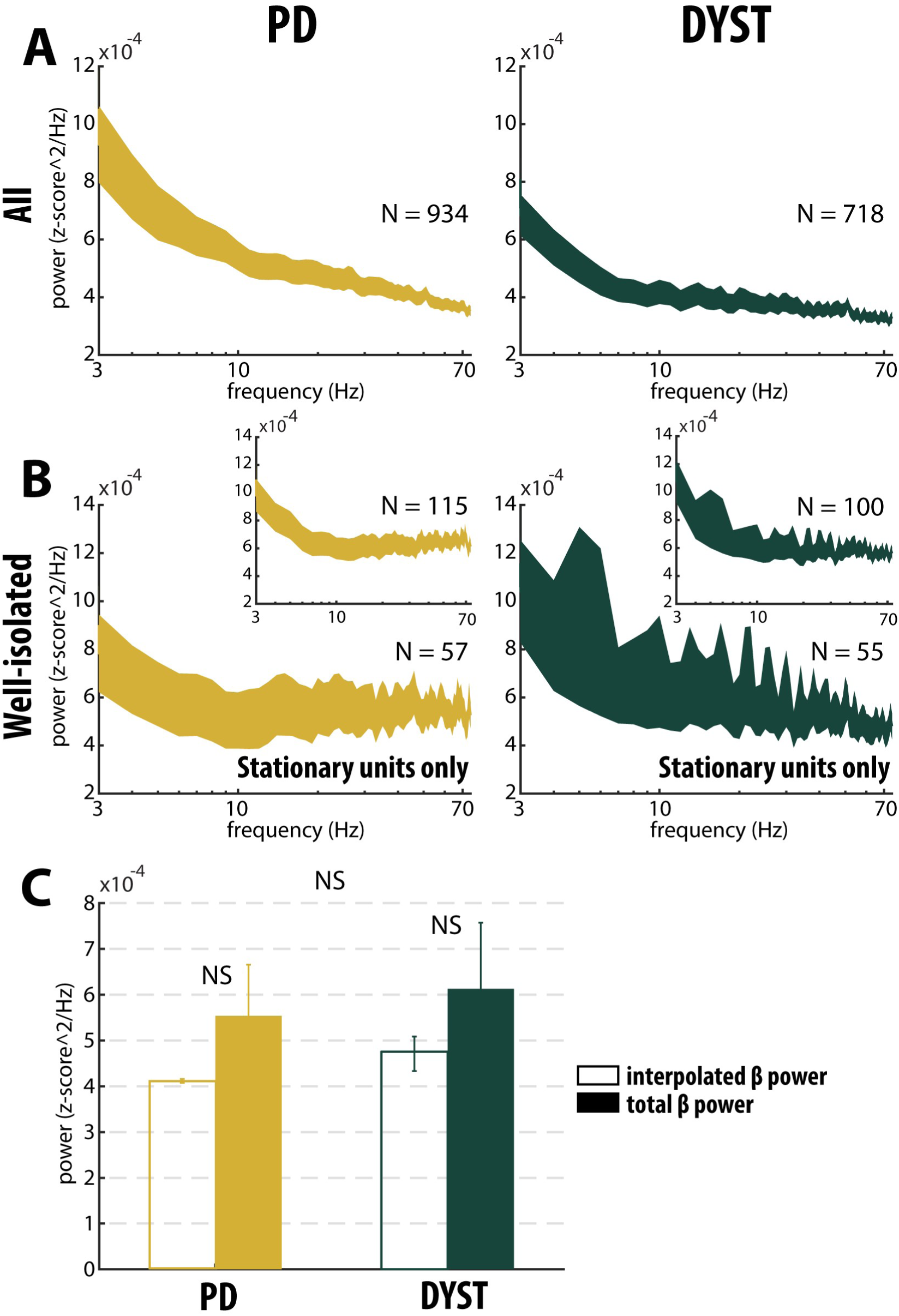
Absence of oscillatory spiking activity in the striatum of parkinsonian (PD) and dystonic (DYST) patients. Average (mean ± SEM) power spectrum densities (PSDs) of all spiking activities **(A)** and only spiking activities recorded in the vicinity of well-isolated stationary units **(B)**. Insets: Average PSDs of the spiking activity recorded in the vicinity of the well-isolated units (non-stationary and stationary units pooled). Abscissas are in log scale. N is the number of spiking activities averaged. **(C)** Average β (13-30Hz) power of the spiking activities recorded in the vicinity of well-isolated stationary units. To interpolate the β power, PSDs were linearly interpolated [based on the two closest points that flanked the 13-30Hz band, namely the values at 12 and 31Hz (spectral resolution of 1Hz)]. Interpolated β power is the mean of the linearly interpolated values between 13 and 30Hz. Total β power is the mean of the observed values between 13 and 30Hz. Error bars represent SEMs. NS: Non significant (Wilcoxon signed rank test and Mann-Whitney U-test).

**Fig.S6.**
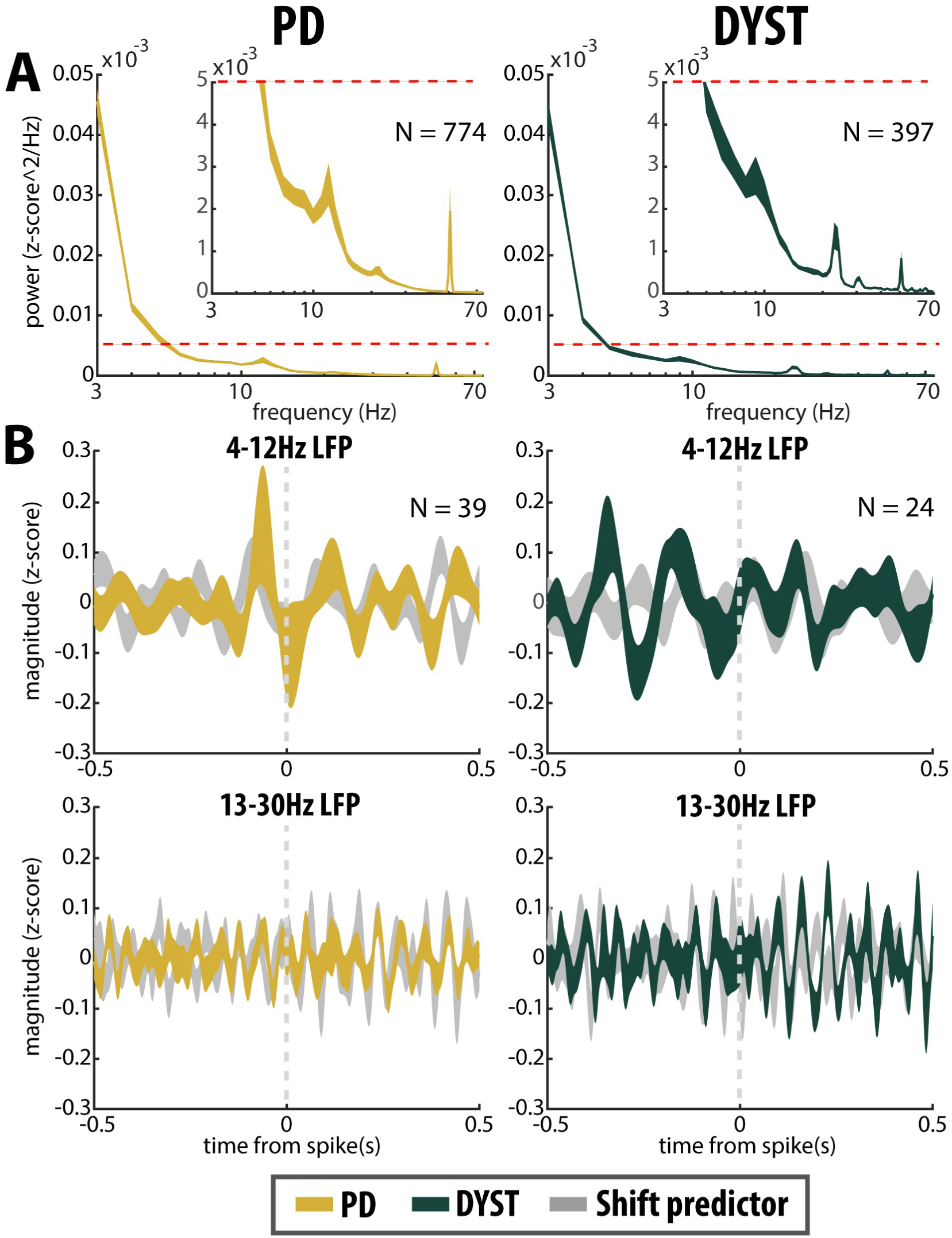
No locking between spike and β LFP oscillations in the striatum of parkinsonian (PD) and dystonic (DYST) patients. **(A)** Average (mean ± SEM) PSDs of striatal LFPs. In the insets, the ordinates are truncated for better visualization of the power. Abscissas are in log scale. N is the number of LFPs averaged. **(B)** Population (mean ± SEM) spike-triggered averages (STAs) of LFP. LFP was recorded in the vicinity of well-isolated stationary units (i.e., spiking activity and LFP were recorded on the same electrode) and offline band-pass filtered from 4 to 12Hz **(upper panels)** or from 13 to 30Hz **(lower panels)**. For comparison, STAs-LFP were also calculated after randomly shifting the timestamp of each spike of the spike train [i.e., random time (comprised between 0 and 1s) was added to the timestamp of each spike of the spike train] in order to abolish any relationship between LFP and spiking activity (Shift predictor). Dashed grey vertical lines indicate the time of the spikes (time = 0). N is the number of STAs-LFP averaged.

**Table S1.** Patient demographic information.

| Patient No. | Disease | Surgery side | Trajectories | Gender | Age at onset | Age at surgery | Disease duration (y) |
| --- | --- | --- | --- | --- | --- | --- | --- |
| 1 | PD | bilateral | R(2) ; L(1) | M | 40 | 62 | 22 |
| 2 | PD | unilateral | R(1) | F | 45 | 62 | 17 |
| 3 | PD | bilateral | R(2) ; L(2) | F | 50 | 62 | 12 |
| 4 | PD | unilateral | L(1) | F | 45 | 62 | 17 |
| 5 | PD | unilateral | R(2) | F | 49 | 60 | 11 |
| 6 | PD | bilateral | R(2) ; L(2) | F | 66 | 72 | 6 |
| 7 | PD | bilateral | R(2) ; L(1) | F | 43 | 59 | 16 |
| 8 | PD | bilateral | R(2) ; L(2) | F | 54 | 68 | 14 |
| 9 | PD | bilateral | R(2) ; L(2) | F | 55 | 62 | 7 |
| 10 | PD | bilateral | R(2) ; L(2) | F | 46 | 58 | 12 |
| 11 | PD | bilateral | R(2) ; L(2) | F | 41 | 57 | 16 |
| 12 | PD | bilateral | R(2) ; L(2) | M | 53 | 66 | 13 |
| 13 | PD | unilateral | L(2) | F | 53 | 63 | 10 |
| 14 | PD | bilateral | R(2) ; L(2) | M | 35 | 44 | 9 |
| 15 | PD | bilateral | R(1) ; L(1) | M | 41 | 57 | 16 |
| 16 | PD | unilateral | R(2) | M | 50 | 57 | 7 |
| 17 | dystonia (NG) | bilateral | R(1) ; L(2) | F | 36 | 56 | 20 |
| 18 | dystonia (NG) | bilateral | R(2) ; L(2) | M | 44 | 49 | 5 |
| 19 | dystonia (NG) | bilateral | R(1) ; L(2) | F | 45 | 65 | 20 |
| 20 | dystonia (NG) | bilateral | R(2) ; L(1) | F | 58 | 60 | 2 |
| 21 | dystonia (NG) | bilateral | R(2) ; L(2) | F | 13 | 19 | 6 |
| 22 | dystonia (NG) | bilateral | R(1) ; L(1) | F | 60 | 63 | 2 |
| 23 | dystonia (NG) | bilateral | R(2) ; L(2) | M | 60 | 62 | 2 |
| 24 | dystonia(NG) | bilateral | R(2) ; L(2) | F | 69 | 71 | 2 |
| 25 | dystonia (G) | bilateral | R(2) ; L(2) | M | 16 | 25 | 9 |
| 26 | dystonia (G) | bilateral | R(2) ; L(2) | M | 12 | 18 | 6 |
| 27 | dystonia (G) | bilateral | R(1) ; L(2) | M | 24 | 39 | 15 |
| 28 | dystonia (G) | bilateral | R(2) ; L(2) | M | 39 | 54 | 15 |
| 29 | dystonia (G) | bilateral | R(1) ; L(2) | M | 53 | 56 | 3 |

PD patients (5 males and 11 females) were $60.7 \pm 6.1$ years old and with a disease duration of $12.8 \pm 4.4$ years (mean ± standard deviation, SD). Dystonic patients (7 males and 6 females) were $49.0.X \pm 18.0$ years old and with a disease duration of $8.2 \pm 6.9$ years (mean ± standard deviation, SD). NG: non-genetic; G: genetic; R: right; L: left; Numbers in parentheses indicate the number of microelectrode trajectories; M: male; F: female.

## References

1. T. Aosaki, M. Miura, T. Suzuki, K. Nishimura, M. Masuda, Acetylcholine-dopamine balance hypothesis in the striatum: an update. Geriatr.Gerontol.Int. 10 Suppl 1, S148–S157 (2010).

2. E. E. Benarroch, Effects of acetylcholine in the striatum. Recent insights and therapeutic implications. Neurology 79, 274–281 (2012).

3. P. Bonsi, et al., Centrality of striatal cholinergic transmission in Basal Ganglia function. Front Neuroanat. 5, 6 (2011).

4. A. Pisani, G. Bernardi, J. Ding, D. J. Surmeier, Re-emergence of striatal cholinergic interneurons in movement disorders. Trends Neurosci. 30, 545–553 (2007).

5. R. Fremont, A. Tewari, C. Angueyra, K. Khodakhah, A role for cerebellum in the hereditary dystonia DYT1. eLife 6, e22775 (2017).

6. R. C. Helmich, The cerebral basis of Parkinsonian tremor: A network perspective. Mov. Disord. 33, 219–231 (2018).

7. R. C. Helmich, M. Hallett, G. Deuschl, I. Toni, B. R. Bloem, Cerebral causes and consequences of parkinsonian resting tremor: a tale of two circuits? Brain 135, 3206–3226 (2012).

8. A. Tewari, R. Fremont, K. Khodakhah, It’s not just the basal ganglia: Cerebellum as a target for dystonia therapeutics. Mov. Disord. 32, 1537–1545 (2017).

9. T. Wu, M. Hallett, The cerebellum in Parkinson’s disease. Brain 136, 696–709 (2013).

10. J. W. Mink, The basal ganglia: focused selection and inhibition of competing motor programs. Prog.Neurobiol. 50, 381–425 (1996).

11. T. Wichmann, Pathophysiologic Basis of Movement Disorders. Prog Neurol Surg 33, 13–24 (2018).

12. P. Limousin, et al., Electrical stimulation of the subthalamic nucleus in advanced Parkinson’s disease. N.Engl.J.Med. 339, 1105–1111 (1998).

13. E. Moro, C. LeReun, J. K. Krauss, A. Albanese, M. Vidailhet, Efficacy of pallidal stimulation in isolated dystonia: a systematic review and meta-analysis. 9 (2017).

14. V. J. Odekerken, et al., GPi vs STN deep brain stimulation for Parkinson disease: Three-year follow-up. Neurology 86, 755–761 (2016).

15. J. L. Ostrem, et al., Subthalamic nucleus deep brain stimulation in isolated dystonia: A 3-year follow-up study. Neurology 88, 25–35 (2017).

16. W. Menegas, et al., Dopamine neurons projecting to the posterior striatum form an anatomically distinct subclass. eLife 4, e10032 (2015).

17. A. C. Bostan, R. P. Dum, P. L. Strick, Cerebellar networks with the cerebral cortex and basal ganglia. Trends Cogn. Sci. (Regul. Ed.) 17, 241–254 (2013).

18. E. Hoshi, L. Tremblay, J. Féger, P. L. Carras, P. L. Strick, The cerebellum communicates with the basal ganglia. Nat. Neurosci. 8, 1491–1493 (2005)

19. R. L. Albin, A. B. Young, J. B. Penney, The functional anatomy of basal ganglia disorders. Trends Neurosci. 12, 366–375 (1989).

20. C. R. Gerfen, et al., D1 and D2 dopamine receptor-regulated gene expression of striatonigral and striatopallidal neurons. Science 250, 1429–1432 (1990).

21. H. Bergman, T. Wichmann, M. R. DeLong, Reversal of experimental parkinsonism by lesions of the subthalamic nucleus. Science 249, 1436–1438 (1990).

22. M. D. Crutcher, M. R. DeLong, Single cell studies of the primate putamen. I. Functional organization. Exp.Brain Res. 53, 233–243 (1984).

23. M. Deffains, E. Legallet, P. Apicella, Modulation of neuronal activity in the monkey putamen associated with changes in the habitual order of sequential movements. J.Neurophysiol. 104, 1355–1369 (2010).

24. M. Kimura, M. Kato, H. Shimazaki, Physiological properties of projection neurons in the monkey striatum to the globus pallidus. Exp.Brain Res. 82, 672–676 (1990).

25. M. Deffains, et al., Subthalamic, not striatal, activity correlates with basal ganglia downstream activity in normal and parkinsonian monkeys. Elife. 5 (2016).

26. M. Filion, L. Tremblay, Abnormal spontaneous activity of globus pallidus neurons in monkeys with MPTP-induced parkinsonism. Brain Res. 547, 142–151 (1991).

27. T. Wichmann, M. R. DeLong, Pathophysiology of Parkinson’s disease: the MPTP primate model of the human disorder. Ann.N.Y.Acad.Sci. 991, 199–213 (2003).

28. D. Guehl, et al., Primate models of dystonia. Progress in Neurobiology 87, 118–131 (2009).

29. K. Kondabolu, et al., Striatal cholinergic interneurons generate beta and gamma oscillations in the corticostriatal circuit and produce motor deficits. Proc Natl Acad Sci U S A 113, E3159–E3168 (2016).

30. N. Lemaire, et al., Effects of dopamine depletion on LFP oscillations in striatum are task- and learning-dependent and selectively reversed by L-DOPA. Proc.Natl.Acad.Sci.U.S.A 109, 18126–18131 (2012).

31. A. Singh, S. M. Papa, Aberrant striatal oscillations after dopamine loss in parkinsonian non-human primates. bioRxiv, 650770 (2019).

32. D. Piña-Fuentes, et al., Toward adaptive deep brain stimulation for dystonia. Neurosurgical Focus 45, E3 (2018).

33. P. Silberstein, et al., Patterning of globus pallidus local field potentials differs between Parkinson’s disease and dystonia. Brain 126, 2597–2608 (2003).

34. M. Deffains, L. Iskhakova, S. Katabi, Z. Israel, H. Bergman, Longer β oscillatory episodes reliably identify pathological subthalamic activity in Parkinsonism. Mov. Disord. (2018) https:/doi.org/10.1002/mds.27418.

35. J. Soares, et al., Role of external pallidal segment in primate parkinsonism: comparison of the effects of 1-methyl-4-phenyl-1,2,3,6-tetrahydropyridine-induced parkinsonism and lesions of the external pallidal segment. J.Neurosci. 24, 6417–6426 (2004).

36. A. A. Kuhn, et al., The relationship between local field potential and neuronal discharge in the subthalamic nucleus of patients with Parkinson’s disease. Exp.Neurol. 194, 212–220 (2005).

37. A. Zaidel, A. Spivak, B. Grieb, H. Bergman, Z. Israel, Subthalamic span of beta oscillations predicts deep brain stimulation efficacy for patients with Parkinson’s disease. Brain 133, 2007–2021 (2010).

38. L. Liang, M. R. DeLong, S. M. Papa, Inversion of dopamine responses in striatal medium spiny neurons and involuntary movements. J.Neurosci. 28, 7537–7547 (2008).

39. A. Singh, et al., Human striatal recordings reveal abnormal discharge of projection neurons in Parkinson’s disease. Proc.Natl.Acad.Sci.U.S.A 113, 9629–9634 (2016).

40. A. Singh, L. Liang, Y. Kaneoke, X. Cao, S. M. Papa, Dopamine regulates distinctively the activity patterns of striatal output neurons in advanced parkinsonian primates. J.Neurophysiol. 113, 1533–1544 (2015).

41. M. Deffains, H. Bergman, Parkinsonism-related β oscillations in the primate basal ganglia networks - Recent advances and clinical implications. Parkinsonism Relat. Disord. (2018) https:/doi.org/10.1016/j.parkreldis.2018.12.015.

42. C. C. Chen, et al., Deep brain stimulation of the subthalamic nucleus: a two-edged sword. Curr.Biol. 16, R952–R953 (2006).

43. N. Mallet, B. Ballion, C. Le Moine, F. Gonon, Cortical inputs and GABA interneurons imbalance projection neurons in the striatum of parkinsonian rats. J. Neurosci. 26, 3875–3884 (2006).

44. A. Sharott, F. Vinciati, K. C. Nakamura, P. J. Magill, A Population of Indirect Pathway Striatal Projection Neurons Is Selectively Entrained to Parkinsonian Beta Oscillations. J.Neurosci. 37, 9977–9998 (2017).

45. L. J. Kish, M. R. Palmer, G. A. Gerhardt, Multiple single-unit recordings in the striatum of freely moving animals: effects of apomorphine and D-amphetamine in normal and unilateral 6-hydroxydopamine-lesioned rats. Brain Res. 833, 58–70 (1999).

46. M. Ketzef, et al., Dopamine Depletion Impairs Bilateral Sensory Processing in the Striatum in a Pathway-Dependent Manner. Neuron 94, 855-865.e5 (2017).

47. J. G. Parker, et al., Diametric neural ensemble dynamics in parkinsonian and dyskinetic states. Nature 557, 177–182 (2018).

48. M. Maltese, J. R. March, A. G. Bashaw, N. X. Tritsch, Dopamine modulates the size of striatal projection neuron ensembles. bioRxiv, 865006 (2019).

49. M. Joshua, S. Elias, O. Levine, H. Bergman, Quantifying the isolation quality of extracellularly recorded action potentials. J.Neurosci.Methods 163, 267–282 (2007).

50. N. Brunel, V. Hakim, Sparsely synchronized neuronal oscillations. Chaos. 18, 015113 (2008).

51. N. Kopell, G. LeMasson, Rhythmogenesis, amplitude modulation, and multiplexing in a cortical architecture. Proc.Natl.Acad.Sci.U.S.A 91, 10586–10590 (1994).

52. Y. Kaneoke, J. L. Vitek, Burst and oscillation as disparate neuronal properties. Journal of Neuroscience Methods 68, 211–223 (1996).

53. M. R. DeLong, Activity of pallidal neurons during movement. J.Neurophysiol. 34, 414–427 (1971).

54. D. Valsky, et al., Real-time machine learning classification of pallidal borders during deep brain stimulation surgery. J. Neural Eng. (2019) https:/doi.org/10.1088/1741-2552/ab53ac (November 5, 2019).

55. M. Deffains, et al., Higher neuronal discharge rate in the motor area of the subthalamic nucleus of Parkinsonian patients. J.Neurophysiol. 112, 1409–1420 (2014).

56. Y. Kawaguchi, C. J. Wilson, S. J. Augood, P. C. Emson, Striatal interneurones: chemical, physiological and morphological characterization. Trends Neurosci. 18, 527–535 (1995).

57. G. Heimer, I. Bar-Gad, J. A. Goldberg, H. Bergman, Dopamine replacement therapy reverses abnormal synchronization of pallidal neurons in the 1-methyl-4-phenyl-1,2,3,6-tetrahydropyridine primate model of parkinsonism. J.Neurosci. 22, 7850–7855 (2002).

58. A. Raz, E. Vaadia, H. Bergman, Firing patterns and correlations of spontaneous discharge of pallidal neurons in the normal and the tremulous 1-methyl-4-phenyl-1,2,3,6-tetrahydropyridine vervet model of parkinsonism. J.Neurosci. 20, 8559–8571 (2000).

59. A. Raz, A. Feingold, V. Zelanskaya, E. Vaadia, H. Bergman, Neuronal synchronization of tonically active neurons in the striatum of normal and parkinsonian primates. J.Neurophysiol. 76, 2083–2088 (1996).

60. S. Hernandez-Lopez, et al., D2 dopamine receptors in striatal medium spiny neurons reduce L-type Ca2+ currents and excitability via a novel PLC[beta]1-IP3-calcineurin-signaling cascade. J. Neurosci. 20, 8987–8995 (2000).

61. S. T. Kitai, D. J. Surmeier, Cholinergic and dopaminergic modulation of potassium conductances in neostriatal neurons. Adv Neurol 60, 40–52 (1993).

62. A. R. West, A. A. Grace, Opposite influences of endogenous dopamine D1 and D2 receptor activation on activity states and electrophysiological properties of striatal neurons: studies combining in vivo intracellular recordings and reverse microdialysis. J. Neurosci. 22, 294–304 (2002).

63. P. Silberstein, Patterning of globus pallidus local field potentials differs between Parkinson’s disease and dystonia. Brain 126, 2597–2608 (2003).

64. G. Buzsaki, C. A. Anastassiou, C. Koch, The origin of extracellular fields and currents--EEG, ECoG, LFP and spikes. Nat.Rev.Neurosci. 13, 407–420 (2012).

65. N. K. Logothetis, The underpinnings of the BOLD functional magnetic resonance imaging signal. J.Neurosci. 23, 3963–3971 (2003).

66. L. Lalla, P. E. Rueda Orozco, M.-T. Jurado-Parras, A. Brovelli, D. Robbe, Local or Not Local: Investigating the Nature of Striatal Theta Oscillations in Behaving Rats. eNeuro 4 (2017).

67. O. Marmor, et al., Local vs. volume conductance activity of field potentials in the human subthalamic nucleus. J Neurophysiol 117, 2140–2151 (2017).

68. A. Machado, et al., Deep brain stimulation for Parkinson’s disease: surgical technique and perioperative management. Mov. Disord. 21 Suppl 14, S247–258 (2006).

69. C. R. Legendy, M. Salcman, Bursts and recurrences of bursts in the spike trains of spontaneously active striate cortex neurons. J.Neurophysiol. 53, 926–939 (1985).

70. A. Moran, H. Bergman, Z. Israel, I. Bar-Gad, Subthalamic nucleus functional organization revealed by parkinsonian neuronal oscillations and synchrony. Brain 131, 3395–3409 (2008).

71. J. A. Goldberg, U. Rokni, T. Boraud, E. Vaadia, H. Bergman, Spike synchronization in the cortex/basal-ganglia networks of Parkinsonian primates reflects global dynamics of the local field potentials. J.Neurosci. 24, 6003–6010 (2004).

